## Appendix for "Point-of-care prognostication in moderate Covid-19: analytical validation and diagnostic accuracy of a soluble urokinase plasminogen activator receptor (suPAR) rapid test"

**SUPPLEMENTARY APPENDIX**

**CONTENTS	PAGE**

1. **STARD checklist**	2
2. **Agreement between RDT and ELISA including samples outside the dynamic range of the RDT**	4
3. **Agreement between repeated RDT measurements**	5
4. **Diagnostic accuracy of the suPAR RDT within the framework of a clinical prediction model**	6

**S1. STARD checklist.**

|  | **Section & Topic** | **No** | **Item** | **Reported on page #** |
| --- | --- | --- | --- | --- |
|  | **TITLE OR ABSTRACT** |  |  |  |
|  |  | **1** | Identification as a study of diagnostic accuracy using at least one measure of accuracy  (such as sensitivity, specificity, predictive values, or AUC) | 1 |
|  | **ABSTRACT** |  |  |  |
|  |  | **2** | Structured summary of study design, methods, results, and conclusions  (for specific guidance, see STARD for Abstracts) | 3 |
|  | **INTRODUCTION** |  |  |  |
|  |  | **3** | Scientific and clinical background, including the intended use and clinical role of the index test | 4 |
|  |  | **4** | Study objectives and hypotheses | 5 |
|  | **METHODS** |  |  |  |
|  | *Study design* | **5** | Whether data collection was planned before the index test and reference standard were performed (prospective study) or after (retrospective study) | 5 |
|  | *Participants* | **6** | Eligibility criteria | 5 |
|  |  | **7** | On what basis potentially eligible participants were identified  (such as symptoms, results from previous tests, inclusion in registry) | 5 |
|  |  | **8** | Where and when potentially eligible participants were identified (setting, location and dates) | 5 |
|  |  | **9** | Whether participants formed a consecutive, random or convenience series | 5 |
|  | *Test methods* | **10a** | Index test, in sufficient detail to allow replication | 6 |
|  |  | **10b** | Reference standard, in sufficient detail to allow replication | 6 |
|  |  | **11** | Rationale for choosing the reference standard (if alternatives exist) | NA |
|  |  | **12a** | Definition of and rationale for test positivity cut-offs or result categories  of the index test, distinguishing pre-specified from exploratory | NA |
|  |  | **12b** | Definition of and rationale for test positivity cut-offs or result categories  of the reference standard, distinguishing pre-specified from exploratory | NA |
|  |  | **13a** | Whether clinical information and reference standard results were available  to the performers/readers of the index test | 6 |
|  |  | **13b** | Whether clinical information and index test results were available  to the assessors of the reference standard | 6 |
|  | *Analysis* | **14** | Methods for estimating or comparing measures of diagnostic accuracy | 6 |
|  |  | **15** | How indeterminate index test or reference standard results were handled | 6 |
|  |  | **16** | How missing data on the index test and reference standard were handled | 8 |
|  |  | **17** | Any analyses of variability in diagnostic accuracy, distinguishing pre-specified from exploratory | 6 |
|  |  | **18** | Intended sample size and how it was determined | 7 |
|  | **RESULTS** |  |  |  |
|  | *Participants* | **19** | Flow of participants, using a diagram | 8 |
|  |  | **20** | Baseline demographic and clinical characteristics of participants | 8 |
|  |  | **21a** | Distribution of severity of disease in those with the target condition | 8 |
|  |  | **21b** | Distribution of alternative diagnoses in those without the target condition | NA |
|  |  | **22** | Time interval and any clinical interventions between index test and reference standard | 5 |
|  | *Test results* | **23** | Cross tabulation of the index test results (or their distribution)  by the results of the reference standard | 8, 9 |
|  |  | **24** | Estimates of diagnostic accuracy and their precision (such as 95% confidence intervals) | 10 |
|  |  | **25** | Any adverse events from performing the index test or the reference standard | NA |
|  | **DISCUSSION** |  |  |  |
|  |  | **26** | Study limitations, including sources of potential bias, statistical uncertainty, and generalisability | 12 |
|  |  | **27** | Implications for practice, including the intended use and clinical role of the index test | 11, 12 |
|  | **OTHER INFORMATION** |  |  |  |
|  |  | **28** | Registration number and name of registry | 7 |
|  |  | **29** | Where the full study protocol can be accessed | 7 |
|  |  | **30** | Sources of funding and other support; role of funders | NA |

**S2. Agreement between the suPAR RDT and ELISA**. Left panel: Bland-Altman plot indicating agreement between the two tests, where samples outside dynamic range of the RDT are set to the limits of detection of the RDT (n = 425). Difference between RDT and ELISA measurement in ng/mL plotted on y-axis. Mean of the RDT and ELISA measurement in ng/mL plotted on x-axis. Limits of agreement defined by the concentrations within which 95% of the data fall. Blue line indicates bias, green line indicates upper limit of agreement, red line indicates lower limit of agreement, all with 95% confidence intervals. Bias = -2.50 ng/mL (95% CI = -2.69 to -2.30), upper limit of agreement = 1.53 ng/mL (95% CI = 1.20 to 1.87), lower limit of agreement = -6.53 ng/mL (95% CI = -6.86 to -6.19). Right panel: Scatterplot indicating correlation between the two tests (R = 0.73 [95% CI = 0.69-0.77]; p < 0.001).

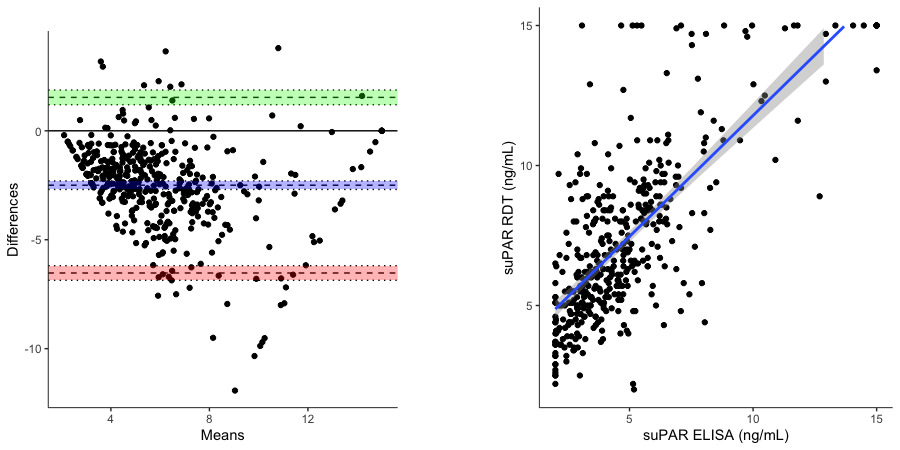

**S3. Agreement between repeated suPAR RDT measurements**. Left panel: Bland-Altman plot indicating agreement between the two RDT measurements (n = 100). Samples outside the dynamic range of the RDT are excluded. Difference between the RDT measurements in ng/mL plotted on y-axis. Mean of the RDT measurements in ng/mL plotted on x-axis. Limits of agreement defined by the concentrations within which 95% of the data fall. Blue line indicates bias, green line indicates upper limit of agreement, red line indicates lower limit of agreement, all with 95% confidence intervals. Bias = 0.50 ng/mL (95% CI = 0.20 to 0.81), upper limit of agreement = 3.52 ng/mL (95% CI = 3.00 to 4.05), lower limit of agreement = -2.52 ng/mL (95% CI = -3.04 to -1.99). Right panel: Scatterplot indicating correlation between the two measurements (R = 0.82 [95% CI = 0.74-0.87]; p < 0.001).

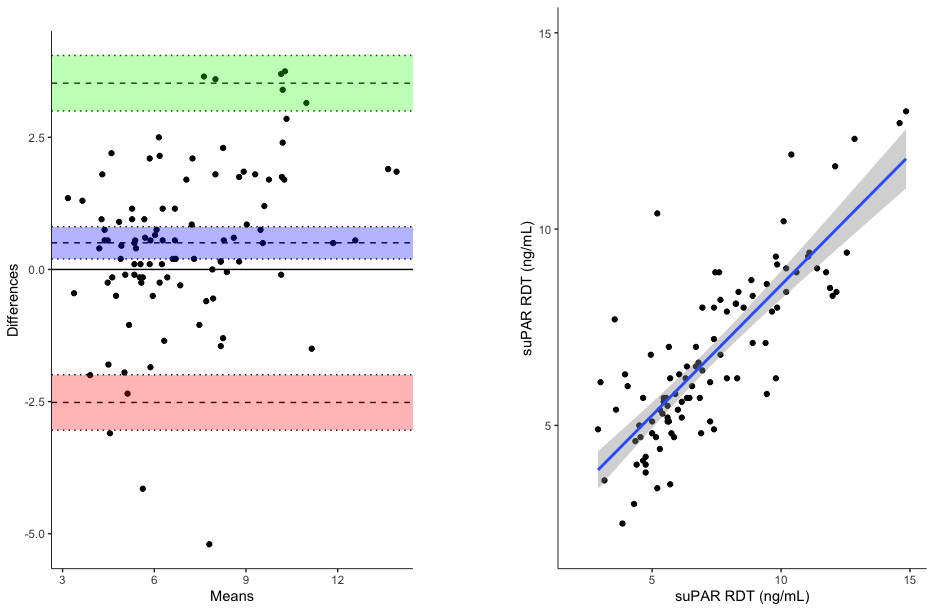

**S4. Diagnostic accuracy the suPAR RDT within the framework of a published clinical prediction model**

***Methods***

Methods for development and validation of the clinical prediction models have been described previously.^1^ Briefly, the cohort was divided into development (n = 256) and external temporal validation (n = 166) cohorts. Linearity between candidate predictors and the primary outcome (supplemental oxygen requirement) was examined using a Lowess smoothing approach and transformations used when serious violations of linearity were detected. Penalised logistic (ridge) regression was used to derive the models in the development cohort. We specified a priori that the models would contain three clinical predictors (age, sex, and baseline SpO_2_) and the biomarker suPAR, measured either using the RDT or ELISA. No variable selection was performed during model development. Model discrimination (c-statistic) and calibration (intercept, slope, and calibration plot) were assessed in the external temporal validation cohort. The clinical utility of the models was assessed using decision curve analyses to compare the net-benefit of triage strategies using the clinical model, the clinical model including suPAR, and an “admit-all” and “admit-none” approach. Predicted classifications were examined across clinically-relevant admission thresholds.

***Results***

Participants who progressed to develop a supplemental oxygen requirement had higher suPAR levels, when quantified using the RDT or ELISA, in both the development and validation cohorts (Table S3.1).

**Table S3.1. Baseline characteristics of development and validation cohorts, stratified by progression to supplemental oxygen requirement.** *Median values (IQR) reported for continuous variables.*

| **Baseline characteristic** | **DEVELOPMENT COHORT** | | | **VALIDATION COHORT** | | |
| --- | --- | --- | --- | --- | --- | --- |
|  | **Overall**  (n = 256) | **Developed oxygen requirement** | | **Overall**  (n = 166) | **Developed oxygen requirement** | |
|  |  | **No**  (n = 206) | **Yes**  (n = 50) |  | **No**  (n = 127) | **Yes**  (n = 39) |
| **Clinical parameters** | | | | | | |
| Age (years) | 52.0  (40.0 to 60.2) | 52.0  (40.0 to 60.0) | 54.0  (42.2 to 62.0) | 54.0  (41.2 to 63.0) | 55.0  (41.5 to 63.0) | 54.0  (41.0 to 66.0) |
| Male sex | 184 / 256  (72%) | 143 / 206  (69%) | 41 / 50  (82%) | 101 / 166  (61%) | 76 / 127  (60%) | 25 / 39  (64%) |
| Oxygen saturation (%) | 98.0  (96.0 to 99.0) | 98.0  (97.0 to 99.0) | 96.0  (95.2 to 98.0) | 98.0  (96.0 to 99.0) | 98.0  (96.0 to 99.0) | 96.0  (95.5 to 98.0) |
| **suPAR concentration** | | | | | | |
| suPAR ELISA (ng/ml) | 4.2  (3.1 to 5.7) | 4.0  (2.9 to 5.5) | 5.4  (4.0 to 6.8) | 4.1  (3.1 to 5.6) | 3.8  (2.9 to 5.1) | 5.5  (3.9 to 6.7) |
| suPAR RDT (ng/ml) | 6.4  (5.1 to 8.8) | 6.0  (4.8 to 8.4) | 8.2  (7.1 to 9.4) | 6.7  (5.2 to 8.7) | 6.2  (5.0 to 8.2) | 8.7  (6.9 to 10.9) |

suPAR concentrations measured on the RDT were transformed prior to model building due to a non-linear relationship with supplemental oxygen requirement. Regression coefficients and adjusted odds ratios for candidate predictors in each model are presented in Table S3.2. After adjustment for the clinical variables, baseline suPAR concentrations were independently associated with development of an oxygen requirement, irrespective of whether they were measured on the RDT or ELISA. The regression coefficient for suPAR in the multivariable prediction model varied depending on which assay was used to quantify suPAR concentrations.

**Table S3.2. Regression coefficients and adjusted associations between candidate predictors and primary outcome in each model in the development cohort**. *^*^ Transformation* $\frac{1}{{({suPAR RDT}/{10)}}^{2}}$ *used due to non-linear relationship between suPAR concentrations and supplemental oxygen requirement. As a result, odds ratios and regression coefficients are not directly comparable. Odds ratios expressed for one unit increase in non-transformed continuous predictors (age, SpO_2_, and suPAR ELISA).*

| **Variable** | **Ridge regression coefficient**  **(95% CI)** | **Adjusted odds ratio**  **(95% CI)** |
| --- | --- | --- |
| **Clinical model** | | |
| Intercept | 42.60  (22.40 to 65.90) | NA |
| Male sex | 0.31  (-0.29 to 1.14) | 1.37  (0.75 to 3.12) |
| Age (years) | 0.02  (-0.02 to 0.02) | 1.00  (0.98 to 1.02) |
| SpO_2_ | -0.46  (-0.69 to -0.24) | **0.63**  **(0.50 to 0.79)** |
| **suPAR ELISA model** | | |
| Intercept | 37.37  (19.63 to 62.53) | NA |
| Male sex | 0.37  (-0.21 to 1.29) | 1.44  (0.81 to 3.63) |
| Age (years) | -0.00  (-0.02 to 0.02) | 1.00  (0.98 to 1.02) |
| SpO_2_ | -0.41  (-0.67 to -0.23) | **0.66**  **(0.51 to 0.80)** |
| suPAR ELISA (mg/l) | 0.14  (0.07 to 0.25) | **1.15**  **(1.07 to 1.29)** |
| **suPAR RDT model** | | |
| Intercept | 40.52  (21.57 to 60.07) | NA |
| Male sex | 0.22  (-0.44 to 1.10) | 1.24  (0.64 to 3.00) |
| Age (years) | -0.01  (-0.03 to 0.01) | 0.99  (0.97 to 1.01) |
| SpO_2_ | -0.42  (-0.62 to -0.23) | **0.66**  **(0.54 to 0.80)** |
| suPAR RDT (ng/ml) ^*^ | -0.37  (-0.53 to -0.22) | **0.69**  **(0.59 to 0.80)** |

Discrimination and calibration of the model containing suPAR was superior to the clinical model and there was no appreciable difference between the ELISA-based and RDT-based models (Figure S3.1).

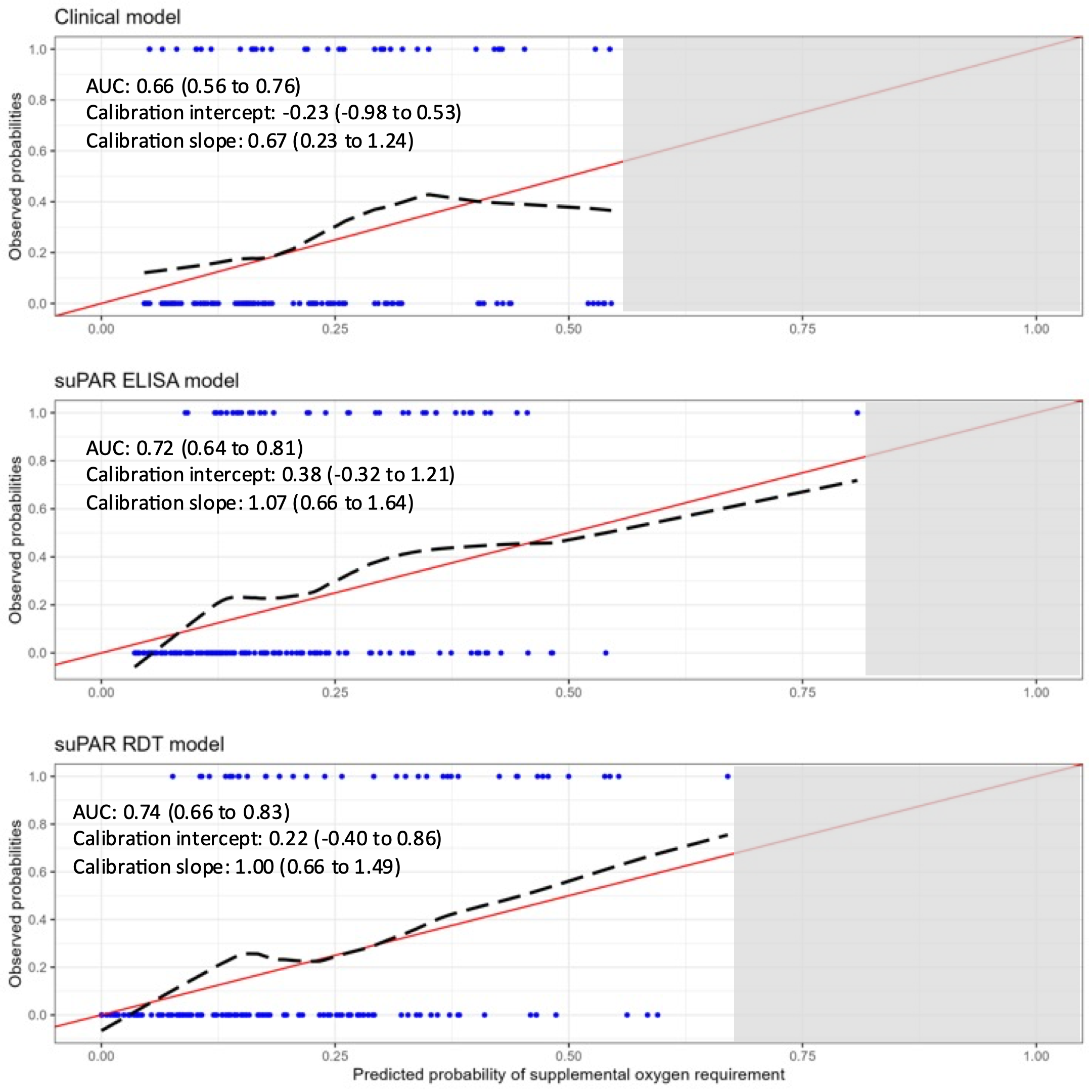
**Figure S3.1. Diagnostic accuracy of clinical prediction models in the validation cohort, with and without suPAR quantified using the RDT or ELISA**. *Red line indicates perfect calibration; black dashed line indicates calibration slope for that particular model; blue rug plots indicate distribution of predicted risk for participants who did (top) and did not (bottom) meet the primary outcome. Transformation used for suPAR measurements on the RDT due to non-linear relationship with the primary outcome. C-statistics indicate how well participants who met the primary outcome are differentiated from those who did not; perfect discrimination is indicated by a c-statistic of 1.0. Calibration slopes indicate agreement between predicted probabilities and observed outcomes; perfect calibration is indicated by a slope of 1.0.*

We recognised that discrimination and calibration are summary measures of model performance and do not necessarily reflect clinical utility. The ability of the models to rule-out progression to oxygen requirement amongst patients presenting with moderate Covid-19 at predicted probabilities (admission thresholds) of 10%, 15%, and 20% is shown (Table S3.3). A cut-off of 10% indicates an approach whereby any patient with a predicted probability of requiring supplemental oxygen ≥ 10% is admitted. The results suggest that both suPAR-containing models were comparable, with the RDT-based model appearing to have slightly greater clinical utility. At a cut-off of 10%, compared to the clinical model, the suPAR-based models would facilitate sending home ~5-10% more patients who would not require supplemental oxygen, whilst simultaneously halving the number of patients discharged who would progress to requiring supplemental oxygen, i.e., an improvement in the ratio of correct to incorrect discharges from 10:1 to 25-30:1.

**Table S3.3. Predicted classification of patients at different cut-offs for each model in the validation cohort.** *A cut-off of 10% reflects a management strategy in which any patient with a predicted risk of requiring oxygen ≥ 10% is admitted. FN = false negative; FP = false positive; TN = true negative; TP = true positive.*

| **Predicted probability of model**  **(cut-off)** | **Per 100 patients (23 patients who would require oxygen)** | | | | **Ratio of incorrect to correct admissions**  **(FP : TP)** | **Ratio of correct to incorrect discharges**  **(TN : FN)** |
| --- | --- | --- | --- | --- | --- | --- |
|  | **Patients who would require oxygen admitted**  **(TP)** | **Unnecessary hospital admissions**  **(FP)** | **Patients who would require oxygen discharged**  **(FN)** | **Patients correctly discharged**  **(TN)** |  |  |
| **Clinical model** |  |  |  |  |  |  |
| 10% | 21 | 56 | 2 | 20 | 3 to 1 | 10 to 1 |
| 15% | 18 | 45 | 5 | 31 | 3 to 1 | 6 to 1 |
| 20% | 14 | 28 | 9 | 48 | 2 to 1 | 5 to 1 |
| **suPAR ELISA model** | | |  |  |  |  |
| 10% | 22 | 52 | 1 | 25 | 2 to 1 | 25 to 1 |
| 15% | 16 | 34 | 7 | 43 | 2 to 1 | 6 to 1 |
| 20% | 13 | 22 | 10 | 54 | 2 to 1 | 5 to 1 |
| **suPAR RDT model** | |  |  |  |  |  |
| 10% | 23 | 46 | 1 | 30 | 2 to 1 | 30 to 1 |
| 15% | 18 | 37 | 5 | 40 | 2 to 1 | 8 to 1 |
| 20% | 14 | 27 | 9 | 50 | 2 to 1 | 6 to 1 |

Decision curve analyses accounting for the differential importance of true positives and false positives (TP and FP; patients admitted who would and would not subsequently require supplemental oxygen) confirmed the superior performance of the model including suPAR and suggested that the RDT-based model may have greater clinical utility (net-benefit) than the model in which suPAR concentrations were quantified using the ELISA (Figure S3.2). When compared to an “admit-all” approach, the clinical model began to add value beyond an admission threshold of 15% (i.e., when the value of a TP is equal to ~7 FPs), whereas the models containing suPAR began to add value at earlier stages (e.g., when bed pressures are less critical), beyond admission thresholds of 5% (ELISA; value of a TP is equal to ~19 FPs) or 2.5% (RDT; value of a TP is equal to ~39 FPs) respectively.

**Figure S3.2. Decision curve analysis for each model in the validation cohort.** *The net benefit for each model is compared to an “admit-all” (red line) and “admit-none” (green line) approach. Transformation used for suPAR measurements on the RDT due to non-linear relationship with the primary outcome. A threshold probability of 5% indicates a scenario where the value of 1 TP (patient admitted who will subsequently require oxygen) is*
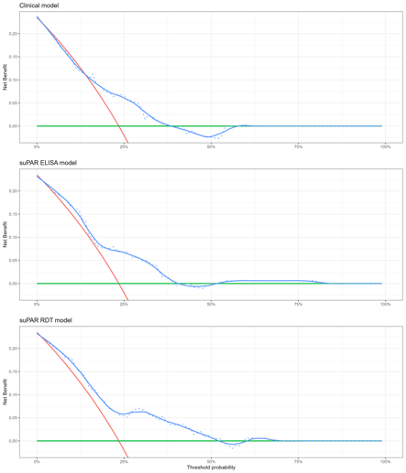
*equivalent to 19 FPs (patients admitted who will not subsequently require oxygen).*
